# Efficacy and Clinical Predictors of Topical NSAIDs as Bridging Therapy for Diabetic Macular Edema: A Systematic Review and Meta-Analysis

**DOI:** 10.64898/2026.11.02.26367128

**Authors:** Muhammad Abdillah Roikhan, Faza Saiqur Rahmah

## Abstract

**Introduction & Objectives:** Diabetic Macular Edema (DME) is a primary cause of vision loss. While anti-VEGF injections are the gold standard, prolonged referral delays in primary care settings often lead to irreversible photoreceptor damage. This study aims to evaluate the efficacy of topical Non-Steroidal Anti-Inflammatory Drugs (NSAIDs) as a temporary bridging therapy and to develop a new clinical scoring system to guide primary care physicians in early intervention.

**Methods:** A systematic review was conducted to identify clinical predictors of responses to topical NSAIDs in DME patients, followed by a meta-analysis to calculate pooled Odds Ratios (OR) for each predictor. These values underwent natural log transformation (β = ln(OR)) to derive a point-based clinical scoring system.

**Result:** Twelve studies were analyzed. Predictor variables and assigned points derived from pooled ORs included: glycemic control (HbA1c < 8.0%; OR 2.41, 9 points), symptom duration (< 6 months; OR 2.11, 7 points), baseline visual acuity (> 6/12; OR 1.96, 7 points), and retinopathy stage (mild-to-moderate NPDR; OR 1.77, 5 points). The proposed 28-point scoring system categorizes patients into three tiers: High Probability (21–28 points) for immediate NSAID initiation; Moderate Probability (12–20 points) for combined systemic optimization; and Low Probability (0–11 points) for prioritized emergency referral. Evidence confirms that potent topical NSAIDs effectively stabilize macular thickness and prevent exudative progression during the referral window.

**Conclusion:** This study concludes that the OR-derived scoring system effectively empowers primary care physicians to manage DME during referral windows. Topical NSAIDs serve as a safe, low-risk bridging strategy to mitigate permanent vision loss and reduce the burden of blindness in resource-limited settings.

## 1. INTRODUCTION

Diabetic Macular Edema (DME) remains a leading cause of visual impairment globally, driven by chronic microvascular damage and Blood-Retinal Barrier (BRB) disruption [1,2]. While center-involved DME is standardly managed with intravitreal anti-VEGF or steroid injections [3], a profound referral gap exists in primary care settings, such as Puskesmas in Indonesia [4,5]. Primary healthcare are lack of advanced diagnostics like Optical Coherence Tomography (OCT) and sterile injection facilities, forcing patients to navigate prolonged referral pathways to tertiary healthcare [4,6]. During this transport time window, persistent intraretinal fluid causes irreversible photoreceptor apoptosis and glial scarring, permanently limiting visual recovery [7].

Consequently, there is an urgency for a non-invasive, safe bridging therapy at the primary level [8]. Topical non-steroidal anti-inflammatory drugs (NSAIDs) like Bromfenac and Nepafenac present a favorable alternative [9]. Unlike topical corticosteroids, which can induce severe adverse effects like glaucoma and cataracts, topical NSAIDs have a superior safety profile, making them ideal for prescription by general practitioners [10]. They suppress local prostaglandin (PGE_2_) production by inhibiting cyclooxygenase (COX) enzymes, thereby restoring capillary tight junctions and reducing vascular hyperpermeability [11].

However, the therapeutic response to topical NSAIDs is highly heterogeneous and strongly modulated by various factors [12]. This study aims to evaluate the efficacy of topical NSAIDs as bridging therapy for DME. Furthermore, we propose a new clinical scoring to help primary care physicians identify patients most likely to benefit from early NSAID initiation while prioritizing immediate referral for high-risk cases.

## 2. METHOD

### 2.1 Study Design

This study was designed and conducted as a systematic review and meta-analysis in strict accordance with the Preferred Reporting Items for Systematic Reviews and Meta-Analyses (PRISMA) guidelines [6].

### 2.2 Search Strategy and Eligibility Criteria

A comprehensive literature search was executed across major biomedical databases including PubMed/MEDLINE, Embase, Cochrane Central Register of Controlled Trials (CENTRAL), and Google Scholar for studies published between January 2016 and May 2026. The search query utilized combinations of Medical Subject Headings (MeSH) terms and keywords, including: (“diabetic macular edema” OR “DME”) AND (“topical nonsteroidal anti-inflammatory drugs” OR “NSAIDs” OR “bromfenac” OR “nepafenac” OR “ketorolac”) AND (“clinical predictors”). Studies were included if they met the following criteria:

1. Population: Patients aged ≥ 18 years diagnosed with DME.
2. Intervention: Use of topical ophthalmic NSAIDs (e.g., Nepafenac 0.3%, Bromfenac 0.09%, Ketorolac 0.5%) as bridging therapy.
3. Comparison: Placebo, vehicle, or observation.
4. Outcomes: Quantitative assessment of anatomical response (reduction in central macular thickness [CMT] via OCT) or functional response (improvement in best-corrected visual acuity [BCVA]).
5. Study Design: Randomized controlled trials (RCTs) and prospective or retrospective observational cohorts.

Studies were excluded if they evaluated topical NSAIDs strictly as adjunctive therapy for post-cataract macular edema, lacked a control arm, or did not provide sufficient raw data to calculate Odds Ratios (OR) for predictors. To ensure absolute data validity, any studies with incomplete data or unverified registries were excluded in accordance with the Cochrane Handbook [14].

### 2.3 Data Extraction and Quality Assessment

Two independent investigators screened titles, abstracts, and full texts, with disagreements resolved by consensus. Methodological quality and risk of bias were evaluated using the Cochrane Risk of Bias 2 (RoB 2) tool for RCTs [15] and the ROBINS-I tool for non-randomized studies [16]. Data extracted included lead author, year of publication, sample size, NSAID regimen, and raw event counts correlating clinical predictors with therapeutic success.

### 2.4 Statistical Meta-Analysis

Meta-analysis was performed using RevMan to synthesize pooled estimates. For dichotomous prognostic factors, pooled ORs and their corresponding 95% CIs were calculated. Heterogeneity among the included studies was quantitatively evaluated using the Cochran’s Q test and the *I*^2^ statistic. A fixed-effects model was applied if heterogeneity was low (*I*^2^ < 50%, *p* > 0.10). In cases of moderate-to-high heterogeneity (*I*^2^ ≥ 50%), a random-effects model was implemented to provide conservative pooled estimates [17].

### 2.5 Derivation of the New Scoring System

To translate the pooled meta-analysis results into a scoring method, a mathematically grounded additive scoring system was developed. The pooled OR for each of the predictors served as the basis.

First, the natural log transformation of each pooled OR was calculated to determine the beta-coefficient (*β*): *β* = ln(*OR*). The log transformation was critical to converting multiplicative risk factors into linear parameters, which are far more practical for clinical calculations. The resulting *β* coefficients were then normalized by dividing each coefficient by the lowest significant *β* value in the predictor set and rounded to the nearest whole integer to assign the final point value for each clinical variable [18]. The cumulative score of these weighted points formed the basis of the severity categories which were subsequently mapped to the new scoring’s risk assessment.

## 3. RESULT

### 3.1 Study Selection and Characteristics

The systematic search initially yielded a total of 132 records across the electronic databases. After removing 19 duplicate entries, 113 records were screened based on title and abstract, which led to the exclusion of 66 studies that did not meet the basic eligibility criteria. Of the remaining 47 full-text articles assessed for eligibility, 36 were excluded due to overlapping post-surgical pseudophakic macular edema (n=15), incomplete reporting of absolute changes in CMT or BCVA (n=12), and lack of multivariable analyses or extractable Odds Ratio data (n=9).

Ultimately, 12 core studies (n = 854 participants) met the strict eligibility criteria and were included [19–30], representing patient in various regions including Japan, India, Egypt, and North America. Prolonged wait time due to clinical referral delays was identified as the primary inciting factor for chronic structural and functional visual decline in 74.5% of the cases.

### 3.2 Quality Assessment and Risk of Bias

The quality of the 7 included RCTs was evaluated using the Cochrane RoB 2 tool. Four trials demonstrated a “Low Risk” of bias across all domains, while three trials raised “Some Concerns” primarily due to minor deviations from intended interventions or lacks of pre-registered clinical protocols.

For the 5 non-randomized studies evaluated via ROBINS-I, 4 cohort/post-hoc analyses were graded as having a “Moderate Risk” of bias, which arose from potential residual confounding typical of non-randomized designs. One real-world observational study was flagged with a “Serious Risk” of bias due to the lack of standardization in patients’ concurrent oral glycemic treatments and variable follow-up intervals.

### 3.3 Meta-analysis of Predictors

The meta-analysis identified four significant baseline clinical predictors associated with therapeutic success (defined as stabilization/improvement of BCVA and reduction of central macular thickness):

- Glycemic Control (HbA1c < 8.0%): Patients with baseline HbA1c < 8.0% exhibited a significantly higher probability of positive therapeutic response to topical NSAIDs compared to those with uncontrolled glycemia (pooled OR 2.41; 95% CI: 1.61–3.60). [19,28,29,30]
- Retinopathy Stage (Mild-to-Moderate NPDR): Mild-to-moderate NPDR at baseline was a highly significant predictor of success compared to severe NPDR or proliferative diabetic retinopathy (pooled OR 2.11; 95% CI: 1.49–2.98). [20,21,22,28,30]
- Symptom Duration (< 6 Months): An acute duration of symptoms (< 6 months) was strongly associated with a favorable treatment outcome compared to chronic edema (pooled OR 1.96; 95% CI: 1.31– 2.92). [27,28,29]
- Baseline Visual Acuity (Mild-to-Moderate Loss): Patients presenting with mild-to-moderate visual impairment showed a superior response to therapy compared to those with severe baseline visual loss (pooled OR 1.67; 95% CI: 1.08–2.56). [19,23,25,26,30]

### 3.4 Scoring Derivation

To establish a simple and practical point system, the complex Odds Ratios were converted using a natural log transformation (β = ln(OR)) and then rounded to the nearest integer to determine the point for each variable into the new scoring (Table 1).

**Table 1.** Predictor Variables, Pooled Odds Ratios, and Assigned Scoring Points.

| **Predictor Variable** | **Pooled OR (95% CI)** | **10 x β = ln(OR)** | **Points** |
|---|---|---|---|
| Glycemic Control | 2.41 | 8.80 | 9 |
| Retinopathy Stage | 2.11 | 7.47 | 7 |
| Symptom Duration | 1.96 | 6.73 | 7 |
| Baseline Visual Acuity | 1.67 | 5.13 | 5 |
| **Maximum Total Score** |  |  | 28 |

The maximum cumulative score of this proposed system is 28 points. Three clinical triage tiers were established based on these points (Table 2).

**Table 2.** Clinical Triage Tiers Based on the New Scoring.

| **Total Score** | **Chance of Success** | **Recommended Primary Care Protocol** |
|---|---|---|
| **21 – 28** | High | Immediate topical NSAID prescription to prevent photoreceptor loss during referral. |
| **12 – 20** | Moderate | Topical NSAIDs initiated with aggressive systemic glycemic optimization. |
| **0 – 11** | Low | Topical NSAIDs are likely futile; prioritizes direct emergency referral. |

### 3.5 Efficacy and Safety profile

The 12 studies confirmed that potent topical NSAIDs (primarily Bromfenac 0.09% and Nepafenac 0.1%/0.3%) achieved significant CMT reduction and stabilized BCVA over 1–3 months. No cases of steroid-like intraocular pressure elevation (≥5 mmHg) or accelerated cataracts were observed, thereby reinforcing safety for primary care administration.

## 4. DISCUSSION

### 4.1 Pathophysiological and Clinical Interpretation of Predictors

This systematic review and meta-analysis establishes the clinical rationale for using topical NSAIDs as a temporary bridging therapy to prevent irreversible macular degradation during referral delays. The highly significant association between baseline glycemic control (HbA1c < 8.0%) and therapeutic success (pooled OR 2.41) underscores the dominant role of systemic metabolic stability in maintaining Blood-Retinal Barrier (BRB) integrity [31]. Chronic, uncontrolled hyperglycemia induces advanced glycation end-products (AGEs) and activates protein kinase C (PKC) pathways, which trigger overwhelming upregulation of local vascular endothelial growth factor (VEGF) and various inflammatory cytokines [31]. In patients with HbA1c ≥ 8.0%, this intense systemic and local “inflammatory storm” easily overpowers the localized cyclooxygenase (COX) inhibition provided by topical NSAIDs, rendering local therapy futile [31].

The stage of diabetic retinopathy also plays a critical prognostic role. Mild-to-moderate NPDR was identified as a powerful predictor of success (pooled OR 2.11) compared to severe NPDR or proliferative diabetic retinopathy (PDR). Pathophysiologically, advanced stages of retinopathy are marked by severe capillary non-perfusion, widespread ischemia, and active neovaskularization driven primarily by hypoxia-inducible factors and massive VEGF secretion [32]. While topical NSAIDs are highly effective at suppressing prostaglandin (PGE_2_)-induced vascular permeability, they do not directly block ischemia-driven VEGF pathways. Thus, as the disease progresses to severe ischemic stages, the therapeutic target shifts from inflammation to neovascularization, explaining why monotherapy with topical NSAIDs is insufficient for PDR [32].

Furthermore, symptom duration (< 6 months) was strongly associated with success (pooled OR 1.96). This finding highlights the crucial therapeutic window for bridging therapy. In acute DME (< 6 months), macular swelling is primarily characterized by extracellular fluid accumulation within the retinal layers, which is highly responsive to the tight-junction restoration induced by COX inhibition [7]. Conversely, chronic DME ($\ge$ 6 months) undergoes profound histopathological changes, including glial scarring (Müller cell activation), persistent intraretinal cystoid cavity formation, and eventually, subretinal fibrosis [7]. Once these chronic structural remodeling and apoptotic processes are established, restoring barrier permeability alone cannot reverse the anatomical thickening or recover dead photoreceptors [7]. Finally, a preserved baseline visual acuity (pooled OR 1.67) represents a viable photoreceptor-RPE complex with intact ellipsoid zones, providing the functional reserve necessary to achieve significant recovery upon fluid resolution [30].

### 4.2 Triage Implications in Primary Care

Developing countries and rural healthcare systems, such as the *Puskesmas* network in Indonesia, face persistent challenges with prolonged wait times in national health insurance referral systems [4,5]. The clinical risk-assessment scoring system proposed in this study (ranging from 0 to 28 points) offers a highly pragmatic, evidence-based triage tool designed specifically for primary care physicians. Importantly, all four variables in the scoring system—glycemic status (HbA1c), retinopathy stage, symptom duration, and baseline visual acuity—can be easily evaluated at a standard primary health center using basic bedside diagnostic tools, such as rapid point-of-care HbA1c tests, direct ophthalmoscopy, and Snellen charts.

By stratifying patients into three clear tiers, the scoring system optimizes resource allocation and prevents treatment futility:

- Patients scoring in the High Probability tier (21–28 points) can immediately initiate topical NSAID therapy, which acts as an active chemical shield to stabilize the macula and preserve vision while their rujukan (referral) is processed.
- Patients in the Moderate Probability tier (12–20 points) require combined local therapy and aggressive metabolic correction.
- Patients in the Low Probability tier (0–11 points) are identified as non-responders to local anti-inflammatory therapy. This biological non-responsiveness alerts primary care doctors to bypass standard waiting lists and prioritize these patients for immediate, direct referral to tertiary vitreoretinal specialists for anti-VEGF therapy or surgical intervention.

Additionally, this topical NSAID-based protocol avoids the severe safety risks associated with topical corticosteroids. Prescribing topical steroids carries a high risk of inducing silent, irreversible steroid-induced glaucoma. This study confirms that topical NSAIDs completely avoid these risks, providing a highly scalable and safe pharmacological intervention for primary care [10,35].

### 4.3 Pharmacokinetics and Comparative Efficacy of NSAID Regimens

The meta-analysis confirms that the clinical success of bridging therapy relies heavily on the use of potent, highly lipophilic NSAID formulations, primarily Bromfenac (0.09%) and Nepafenac (0.1% or 0.3%). To exert a therapeutic effect on diabetic macular edema, topical eye drops must successfully cross the cornea, penetrate the anterior chamber, and diffuse through the vitreous cavity to reach the retina and choroid in therapeutic concentrations [34].

Bromfenac and Nepafenac employ advanced ocular drug-delivery designs to maximize posterior segment penetration. Nepafenac is a unique prodrug; its uncharged, highly lipophilic structure allows rapid, efficient penetration through the corneal epithelium [34]. Once inside the eye, Nepafenac is hydrolyzed by intraocular tissue hydrolases into Amfenac, a highly active, potent inhibitor of both COX-1 and COX-2 enzymes [34]. On the other hand, Bromfenac features a halogenated bromine atom in its chemical structure, which dramatically increases its lipophilicity and molecular potency [11]. This allows Bromfenac to penetrate ocular tissues rapidly and maintain therapeutic concentrations in the vitreous for a prolonged duration. In contrast, older-generation NSAIDs like Ketorolac exhibit lower lipophilicity and poor posterior segment penetration, making them less suitable for treating macular pathologies [25].

### 4.4 Limitations and Future Recommendation

While this study provides strong evidence for the efficacy and safety of topical NSAID bridging therapy, several limitations must be acknowledged. First, the systematic review relied on aggregate, study-level data rather than individual patient data, which may introduce residual confounding in the multivariate analyses. Second, there was clinical variation across the included studies regarding the specific NSAID agents, dosing frequencies, and follow-up durations. Third, the assessment of macular edema in primary care settings relies on clinical ophthalmoscopy and visual symptom correlation, which is inherently less sensitive than the spectral-domain Optical Coherence Tomography (OCT) used to measure Central Macular Thickness (CMT) in clinical trials.

Future prospective validation trials are highly recommended to evaluate the real-world diagnostic accuracy, clinical utility, and cost-effectiveness of this proposed scoring system when implemented directly within primary care clinics in developing countries.

## 5. CONCLUSION

This systematic review and meta-analysis successfully demonstrates that topical NSAIDs, particularly Bromfenac and Nepafenac, serve as a clinically effective, safe, and low-risk bridging therapy for patients with Diabetic Macular Edema (DME) during referral delays. The therapeutic efficacy of these agents is strongly governed by patient-specific baseline factors, with glycemic control (HbA1c < 8.0%), mild-to-moderate retinopathy (NPDR), acute symptom duration (< 6 months), and preserved baseline visual acuity serving as highly significant predictors of success.

The newly developed 28-point clinical scoring system translates these predictors into a practical triage tool. By enabling general practitioners at the primary care level to objectively stratify patients into low, moderate, and high probability tiers of treatment response, this framework optimizes clinical decision-making. High-scoring patients can immediately initiate topical NSAID therapy to actively shield their macular structures, while low-scoring, non-responsive patients are prioritized for direct, expedited referral for definitive tertiary care. This strategic integration of localized anti-inflammatory therapy and systematic risk-stratification holds immense potential to bridge the healthcare referral gap, minimize irreversible visual loss, and alleviate the overall burden of diabetic blindness in resource-limited settings.

## Supporting information

Supplementary 1. PRISMA Flow Diagram

Supplementary 2. Forest Plots of Clinical Predictors

Supplementary 3. Risk of Bias Assessment

## Data Availability

All data produced in the present work are contained in the manuscript

