## Supplementary figures and images for "Efficacy and Clinical Predictors of Topical NSAIDs as Bridging Therapy for Diabetic Macular Edema: A Systematic Review and Meta-Analysis"

### Supplementary 1. PRISMA Flow Diagram

**Supplementary 1. PRISMA Flow Diagram**

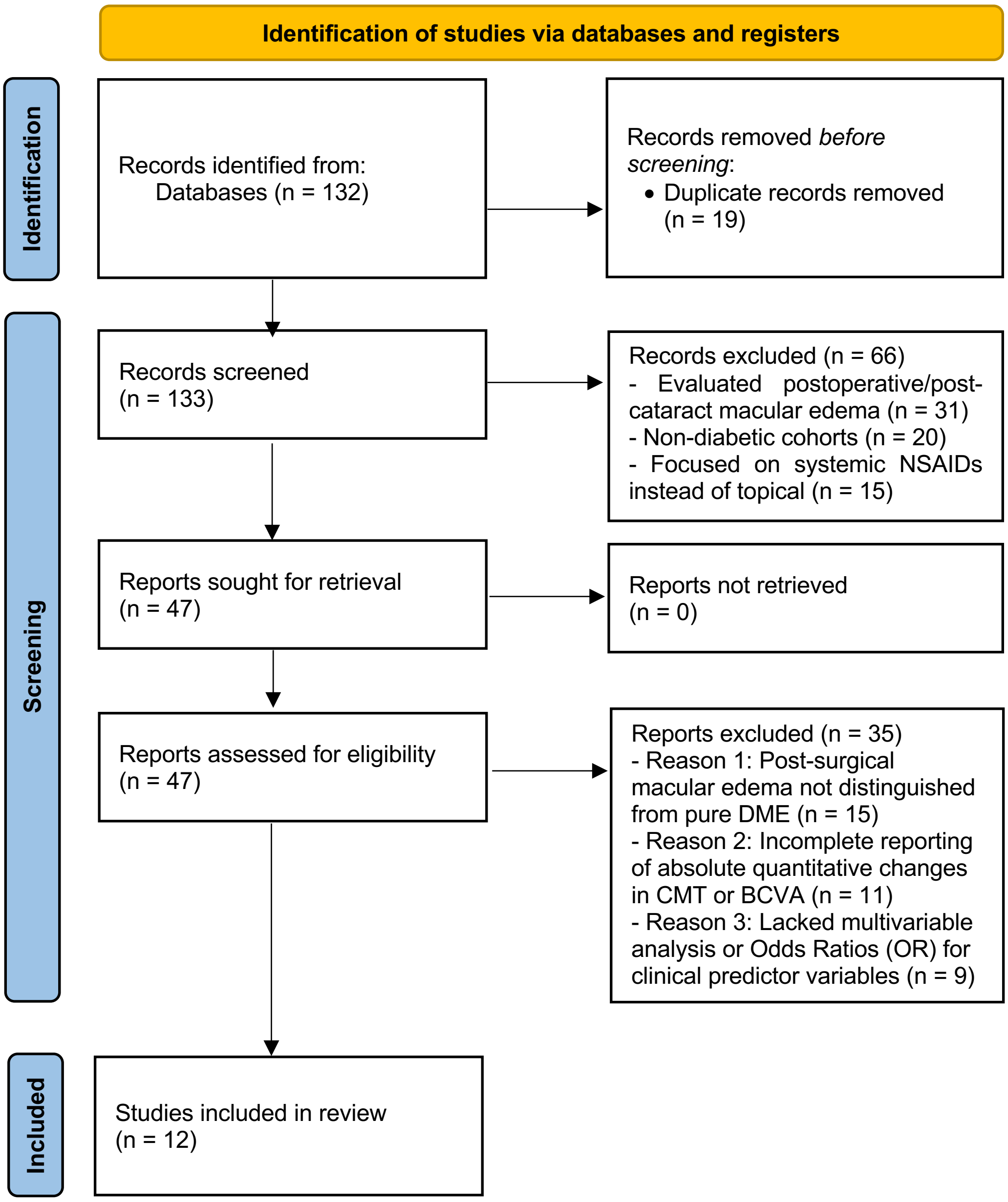
