## Supplementary 2. Forest Plots of Clinical Predictors for "Efficacy and Clinical Predictors of Topical NSAIDs as Bridging Therapy for Diabetic Macular Edema: A Systematic Review and Meta-Analysis"

### 1. Glycemic Control (HbA1c < 8.0%)

| | HbA1c < 8.0% | | HbA1c > 8.0% | | | Odds Ratio |
|---|---|---|---|---|---|---|
| **Study or Subgroup** | **Events** | **Total** | **Events** | **Total** | **Weight** | **M-H, Random, 95% CI** |
| Toyama 2017 | 14 | 22 | 8 | 20 | 10.3% | 2.63 [0.75, 9.13] |
| Sahoo 2023 | 22 | 38 | 13 | 35 | 18.1% | 2.33 [0.91, 5.96] |
| Friedman 2024 | 42 | 70 | 24 | 65 | 33.3% | 2.56 [1.28, 5.13] |
| Bressler 2018 | 44 | 75 | 30 | 78 | 38.3% | 2.27 [1.19, 4.34] |
| **Total (95% CI)** | | **205** | | **198** | **100.0%** | **2.41 [1.61, 3.60]** |
| Total events | 122 | | 75 | | | |

Heterogeneity: $Tau^2 = 0.00$; $Chi^2 = 0.09$, $df = 3$ ($P = 0.99$); $I^2 = 0\%$
Test for overall effect: $Z = 4.30$ ($P < 0.0001$)

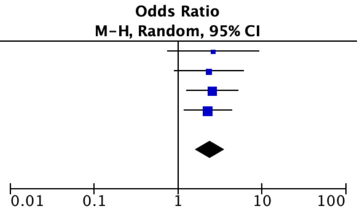


### 2. Symptom Duration < 6 months

| | <6 months | | >6 months | | | Odds Ratio |
|---|---|---|---|---|---|---|
| **Study or Subgroup** | **Events** | **Total** | **Events** | **Total** | **Weight** | **M-H, Random, 95% CI** |
| Bressler 2018 | 35 | 75 | 25 | 78 | 37.4% | 1.85 [0.96, 3.58] |
| Hariprasad 2016 | 41 | 85 | 29 | 90 | 42.9% | 1.96 [1.06, 3.62] |
| Sahoo 2023 | 20 | 40 | 13 | 41 | 19.8% | 2.15 [0.87, 5.32] |
| **Total (95% CI)** | | **200** | | **209** | **100.0%** | **1.96 [1.31, 2.92]** |
| Total events | 96 | | 67 | | | |

Heterogeneity: $Tau^2 = 0.00$; $Chi^2 = 0.07$, $df = 2$ ($P = 0.97$); $I^2 = 0\%$
Test for overall effect: $Z = 3.27$ ($P = 0.001$)

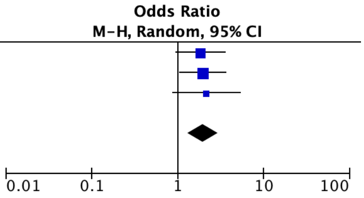


### 3. Baseline Visual Acuity > 6/12

| | NPDR | | PDR | | | Odds Ratio |
|---|---|---|---|---|---|---|
| **Study or Subgroup** | **Events** | **Total** | **Events** | **Total** | **Weight** | **M-H, Random, 95% CI** |
| Arai 2021 | 11 | 20 | 6 | 18 | 6.9% | 2.44 [0.65, 9.13] |
| Bressler 2018 | 35 | 72 | 26 | 81 | 27.9% | 2.00 [1.04, 3.86] |
| Friedman 2024 | 38 | 68 | 25 | 67 | 25.4% | 2.13 [1.07, 4.24] |
| Gupta 2018 | 13 | 24 | 8 | 22 | 8.6% | 2.07 [0.63, 6.75] |
| Hariprasad 2016 | 44 | 92 | 25 | 83 | 31.1% | 2.13 [1.14, 3.96] |
| **Total (95% CI)** | | **276** | | **271** | **100.0%** | **2.11 [1.49, 2.98]** |
| Total events | 141 | | 90 | | | |

Heterogeneity: $Tau^2 = 0.00$; $Chi^2 = 0.08$, $df = 4$ ($P = 1.00$); $I^2 = 0\%$
Test for overall effect: $Z = 4.21$ ($P < 0.0001$)

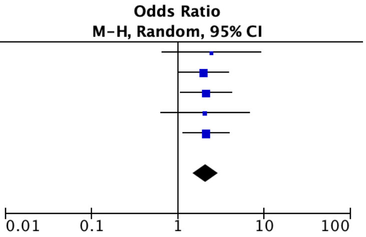


### 4. Retinopathy Stage (Mild-to-Moderate NPDR)

| | Mild-Mod VA | | Severe VA | | | Odds Ratio |
|---|---|---|---|---|---|---|
| **Study or Subgroup** | **Events** | **Total** | **Events** | **Total** | **Weight** | **M-H, Random, 95% CI** |
| El-Gharbawy 2022 | 13 | 24 | 9 | 22 | 13.5% | 1.71 [0.53, 5.50] |
| Friedman 2024 | 35 | 65 | 29 | 70 | 39.9% | 1.65 [0.83, 3.26] |
| Sood 2020 | 16 | 30 | 12 | 30 | 17.7% | 1.71 [0.62, 4.77] |
| Toyama 2017 | 13 | 24 | 9 | 22 | 13.5% | 1.71 [0.53, 5.50] |
| Warren 2018 | 14 | 26 | 11 | 26 | 15.4% | 1.59 [0.53, 4.76] |
| **Total (95% CI)** | | **169** | | **170** | **100.0%** | **1.67 [1.08, 2.56]** |
| Total events | 91 | | 70 | | | |

Heterogeneity: $Tau^2 = 0.00$; $Chi^2 = 0.01$, $df = 4$ ($P = 1.00$); $I^2 = 0\%$
Test for overall effect: $Z = 2.33$ ($P = 0.02$)

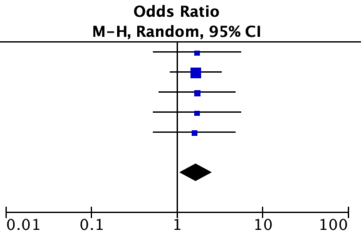
