## Supplementary 3. Risk of Bias Assessment for "Efficacy and Clinical Predictors of Topical NSAIDs as Bridging Therapy for Diabetic Macular Edema: A Systematic Review and Meta-Analysis"

Two standardized, evidence-based tools were utilized based on study design:

1. **Cochrane Risk of Bias 2 (RoB 2) Tool** for Randomized Controlled Trials (RCTs).
2. **ROBINS-I Tool** (Risk of Bias in Non-randomized Studies of Interventions) for non-randomized clinical trials, prospective comparative studies, observational cohorts, and post-hoc analyses.

Section 1: Randomized Controlled Trials (RoB 2 Assessment)

The Cochrane RoB 2 tool evaluates five distinct domains of potential bias:

- D1: Bias arising from the randomization process
- D2: Bias due to deviations from intended interventions
- D3: Bias due to missing outcome data
- D4: Bias in measurement of the outcome
- D5: Bias in selection of the reported result
- Overall: Summary risk of bias rating

Table 1: Summary of Cochrane RoB 2

| **Study ID** | **D1** | **D2** | **D3** | **D4** | **D5** | **Overall Bias** |
|---|---|---|---|---|---|---|
| Hariprasad 2016 | 🟢 | 🟢 | 🟢 | 🟢 | 🟢 | Low Risk (🟢) |
| Khurana 2016 | 🟢 | 🟡 | 🟢 | 🟢 | 🟢 | Some Concerns (🟡) |
| Toyama 2017 | 🟢 | 🟢 | 🟢 | 🟢 | 🟢 | Low Risk (🟢) |
| Warren 2018 | 🟢 | 🟡 | 🟢 | 🟢 | 🟡 | Some Concerns (🟡) |
| Radwan 2019 | 🟡 | 🟢 | 🟢 | 🟡 | 🟢 | Some Concerns (🟡) |
| Sood 2020 | 🟢 | 🟢 | 🟢 | 🟢 | 🟢 | Low Risk (🟢) |
| El-Gharbawy 2022 | 🟢 | 🟢 | 🟢 | 🟢 | 🟢 | Low Risk (🟢) |

*Ratings: Low Risk (🟢), Some Concerns (🟡), High Risk (🔴)*

Study-by-Study Justifications:

1. Hariprasad 2016

   Randomization was computer-generated with central allocation concealment. The double-masked design (placebo vs. active drug) was strictly maintained. High retention rate and pre-registered trial protocols resulted in an overall Low Risk rating.
2. Khurana 2016

   Clear randomization and blinding of patients were reported. However, some concerns were noted in Domain 2 (deviations from intended interventions) due to minor compliance tracking challenges associated with at-home topical drop administration.
3. Toyama 2017

   This prospective, randomized, double-masked clinical trial utilized solid allocation concealment methods. Dropouts were thoroughly accounted for using intent-to-treat (ITT) analysis. Visual and anatomical outcomes were measured objectively via standard protocols, leading to an overall Low Risk rating.
4. Warren 2018

   While random sequence generation was clear, some concerns were flagged in Domain 5 because the study served as a small pilot trial, and some secondary exploratory outcomes were not predefined in a public registry before trial commencement.

5. Radwan 2019

   The study did not explicitly detail the method used for random sequence generation and allocation concealment, leading to "Some Concerns" in Domain 1. Additionally, masking details of the outcome assessors were sparse (Domain 4).

6. Sood 2020

   Well-executed comparative trial. Adequate randomization, masking of investigators, and complete outcome data collection with zero lost-to-follow-up in either active NSAID arm.

7. El-Gharbawy 2022

   Solid trial methodology with low attrition rates. Randomization was robust, and outcome measurement (macular thickness changes via spectral-domain OCT) was fully standardized and blinded to the assessors.

<u>Section 2: Non-Randomized Studies of Interventions (ROBINS-I Assessment)</u>

The ROBINS-I tool assesses risk of bias across seven domains:

- D1: Bias due to confounding
- D2: Bias in selection of participants into the study
- D3: Bias in classification of interventions
- D4: Bias due to deviations from intended interventions
- D5: Bias due to missing data
- D6: Bias in measurement of outcomes
- D7: Bias in selection of the reported result
- Overall: Summary rating

Table 2: Summary of ROBINS-I

| **Study ID** | **D1** | **D2** | **D3** | **D4** | **D5** | **D6** | **D7** | **Overall Bias** |
|---|---|---|---|---|---|---|---|---|
| Bressler 2018 | 🟡 | 🟢 | 🟢 | 🟢 | 🟢 | 🟢 | 🟢 | Moderate (🟡) |
| Gupta 2018 | 🟡 | 🟢 | 🟢 | 🟢 | 🟡 | 🟢 | 🟢 | Moderate (🟡) |
| Arai 2021 | 🟡 | 🟡 | 🟢 | 🟢 | 🟢 | 🟢 | 🟢 | Moderate (🟡) |
| Sahoo 2023 | 🔴 | 🟡 | 🟢 | 🟢 | 🟢 | 🟢 | 🟢 | Serious (🔴) |
| Friedman 2024 | 🟡 | 🟢 | 🟢 | 🟢 | 🟢 | 🟢 | 🟢 | Moderate (🟡) |

*Ratings: Low Risk (🟢), Moderate Risk (🟡), Serious Risk (🔴)*

Study-by-Study Justifications:

1. Bressler 2018

   Since this was a post-hoc analysis of a randomized cohort, the baseline characteristics were well-controlled. However, because it was retrospective in nature and not originally powered for the post-hoc sub-groups, a baseline risk of residual confounding (Domain 1) remains, leading to an overall Moderate Risk rating.

2. Gupta 2018

   This non-randomized prospective trial comparing Bromfenac vs. Nepafenac had minimal missing data. However, there was a minor potential for confounding (Domain 1) as treatment allocation was determined by clinical logistics rather than random selection.

3. Arai 2021 (Prospective cohort)

   This study lacked a randomized design, creating moderate risk of confounding. Baseline glycemic statuses and retinopathy severities varied slightly across patients. Nonetheless, the prospective design, clear classification of intervention, and robust OCT-based anatomical measurements kept the overall bias to Moderate.

4. Sahoo 2023

This real-world prospective observational study carried a Serious Risk of confounding (Domain 1). Because patient enrollment reflected real-world clinical practice, there was no control over systemic treatments (such as varying oral hypoglycemic drugs) or follow-up intervals, which introduces significant confounders for visual recovery.

5. Friedman 2024

Although the cohort was rigorously monitored, the retrospective nature of identifying prognostic biomarkers retrospectively introduces moderate selection and confounding concerns (Domain 1). Standardized OCT measurements minimized outcome measurement bias.
